# Pharmacological vs non-pharmacological treatment in the management of Relative Energy Deficiency in Sport (RED-S): A systematic review and meta-analysis

**DOI:** 10.1101/2025.07.18.25331767

**Authors:** A Wood, A Soundy

## Abstract

**Objective:** To conduct a systematic review assessing the impact of pharmacological and non-pharmacological interventions on Relative Energy Deficiency in Sport (RED-S).

**Design:** Systematic review and meta-analysis

Data source CINHAL, MEDLINE, SportDiscus, ERIC, Embase from inception until July 2025.

Eligibility criteria for selecting studies Experimental, quasi and pre-experimental literature that investigated interventions designed to support the symptoms of RED-S.

**Results:** A total of 19 studies (15 non-pharmacological interventions, 4 pharmacological interventions) were included in the review. Non-pharmacological interventions demonstrated positive benefits on menstrual function recovery, energy availability, fat mass and body fat percentage. Pharmacological interventions demonstrated positive results for biomarkers. Non-pharmacological Interventions were limited by quality.

**Conclusion:** Current evidence base favours non-pharmacological management as an initial response for managing RED-S. Initial pharmacological management appears appropriate in specific situations. Further research is needed to help develop understanding.

**What is already known on this topic:**

- Evidence is limited, with few high quality randomised controlled trials comparing treatment options.
- Management of RED-S prioritises non-pharmacological approaches, mainly nutritional rehabilitation, training load adjustments, and education to address LEA. With pharmacological treatments reserved for persistent or severe cases.

**What this study adds:**

- Non-pharmacological interventions identified moderate to high effect size improvements on menstrual function recovery and energy availability. However, evidence was supported by low confidence from the GRADE assessment.
- Non-pharmacological interventions demonstrated moderate to very high effect size, supported by meta-analysis, for body composition measures such as fat mass and fat percentage. Evidence was supported with moderate confidence from the GRADE assessment.
- Both pharmacological and non-pharmacological studies targeted hormonal changes and biomarkers. Non-pharmacological studies identified low confidence in evidence, despite demonstrating moderate to high effect size with meta-analysis, showing further considerations are needed for hormonal changes and biomarkers. Pharmacological studies identified moderate confidence in evidence showing in moderate effect size evidence for BMD markers and hormonal profiling, but low confidence in clinically meaningful effects of changes in bone turnover markers.

**How this study might affect research, practice or policy:**

- Findings from the current review apply to female groups of athletes primarily engaged in distance or endurance-based sports. Most studies were conducted in university settings, and the most common type of intervention involved dietary adaptations, delivered in 80% of interventions by a dietitian or expert in nutrition. These parameters should be considered within clinical recommendations given and taken into consideration for future research, practice and policy.
- The development and adoption of standardised definitions and core outcomes are urgently needed to ensure constancy and comparability across studies.
- Future trials should investigate the combined use of non-pharmacological and pharmacological strategies, aiming to clarify optimal treatment sequencing, potential synergistic effects, and personalised approaches to RED-S management.

## Introduction

Relative Energy Deficiency in Sport (RED-S) a relatively new and evolving clinical model that is defined as “*A syndrome of impaired physiological and/or psychological functioning experienced by female and male athletes that is caused by exposure to problematic (prolonged and/or severe) low energy availability (LEA). The detrimental outcomes include, but are not limited to, decreases in energy metabolism, reproductive function, musculoskeletal health, immunity, glycogen synthesis and cardiovascular and haematological health, which can all individually and synergistically lead to impaired well-being, increased injury risk and decreased sports performance*”^1^. LEA results when the energy intake of an athlete is insufficient to meet the demands of both exercise and normal physiological function^2^. The term RED-S stems from and the earlier term "Female Athlete Triad" (initially defined in 1993^3^ as the interrelationship between disordered eating, amenorrhoea, and osteoporosis). RED-S recognizes a broader range of health and performance consequences in both female and male athletes. This syndrome is associated with a wide array of clinical outcomes, including menstrual dysfunction, impaired bone health, compromised immune function, gastrointestinal disturbances, fatigue, reduced cardiovascular and metabolic function, and poor mental health^4,5^. Contributing factors to RED-S are multifactorial and may include excessive training, inadequate nutrition, mental health challenges, disordered eating or eating disorders, poor sleep, illness, or undiagnosed medical conditions^2^. RED-S is highly prevalent in elite sport settings; for instance, studies report signs of LEA in both male volleyball players and professional female football players, with many athletes at risk of developing the full spectrum of RED-S^6,7^. Around 70% of elite athletes have a medium to high risk of RED-S^8^, but studies have also shown a higher prevalence of symptoms. For instance, an Australian study found that 80% of elite and pre-elite athletes exhibited at least one RED-S-related symptom, and 37% presented with two or more^9^. Prevalence is not only applicable in a sports setting, but also clinical as Functional Hypothalamic Amennorhea (FHA) accounts for 20-35% of secondary amenorrhea cases^10^.

The widespread prevalence and serious health implications of RED-S underscore the importance of effective management strategies, both pharmacological and non-pharmacological, that have been identified in athletic populations^2^ as such strategies help mitigate the impact of RED-S on athletes^1^. Currently, non-pharmacological treatment forms the cornerstone of RED-S management, with an emphasis on addressing the underlying issue of LEA. These strategies primarily involve educational initiatives aimed at increasing awareness and understanding of RED-S among athletes, coaches, and healthcare professionals (a primary intervention strategy^11^). In addition, tertiary prevention strategies centred around individualized nutritional interventions are critical and show promising effect^11^, focusing on increasing overall energy intake to restore energy balance and support physiological recovery^12^. Modifications to training load, either by reducing volume or intensity, are also commonly implemented to decrease Energy Expenditure (EE) and allow for physiological restoration^13^. These non-pharmacological approaches are often effective in reversing early symptoms of RED-S and remain the first-line treatment, particularly in the absence of severe clinical manifestations. Where non-pharmacological modalities of treatment are ineffective (notably in particular groups like females with resistant amenorrhoea or with low bone mineral density (BMD)), not appropriate, or when physiological effects mean immediate intervention is required, pharmacological strategies can be considered^14^. Supportive treatments, such as calcium, vitamin D, and iron supplementation, are often implemented as part of a broader therapeutic strategy^11,15^. Although more specific strategies have been tested and utilised. For instance, transdermal 17β-oestradiol with cyclic progesterone has been shown to be more effective than combined oral contraceptives (COCs) in improving BMD in oligo-amenorrhoeic athletes^16^. Further to this, treatments like hormone replacement or bone-active agents may target complications such as low bone density when conservative measures fail^16,17^. Treatments such as these can be considered controversial due to long-term safety concerns^18^ and further research is required to clarify their role in RED-S management.

Recent reviews, including those by Tenforde et al.^19^ and Melin et al.^16^, provide important insights into the pathophysiology and management of RED-S. However, they largely focus on descriptive frameworks rather than interventional comparisons, and there is a noticeable lack of high-quality comparative evidence on treatment efficacy. The International Olympic Committee’s (IOC) narrative review on primary, secondary, and tertiary prevention strategies underscores the importance of addressing LEA as the root cause, recommending tailored nutritional rehabilitation, training adjustments, and multidisciplinary care^11^. Past reviews consistently call for more evidence-based guidelines and emphasize that most pharmacological options address secondary outcomes (e.g., menstrual restoration or bone health) rather than the underlying LEA. To date, no systematic review or meta-analysis has directly compared the efficacy, risks, and outcomes of pharmacological versus non-pharmacological treatments for RED-S. This gap is critical given the rising prevalence of RED-S and the growing diversity of affected athletic populations. Thus, the present systematic review and meta-analysis aims to fill this gap by rigorously evaluating available interventions for RED-S.

## Methods

A systematic review was undertaken and reported according to the PRISMA checklist^20^. A protocol was developed and published on the 17 June 2025 on PROSPERO (reference: CRD420251073240).

### Eligibility criteria

Studies were included if they met the following conditions as set out by the acronym PICOS.

#### Participants

To be included studies need to include female athletes classified as elite, recreational, pre-elite, club, active or other level. All athletes had to have characteristics identified within the remit of REDs^1^ . Studies were included if they identified participants with oligo-amenorrhea, LEA, FHA, and/or exercise-related menstrual dysfunction (ExMD). Studies were excluded if participants were male, had a clinical diagnosis of a psychological disorder or mental illness, were currently using hormonal contraceptives, or had Polycystic Ovary Syndrome (PCOS) or Endometriosis. Studies using the same group of participants without assessing different outcome measures were excluded.

#### Intervention

Interventions were grouped as pharmacological or non-pharmacological interventions. Pharmacological interventions were included if they utilised Oestrogen therapy, Hormone Replacement Therapy (HRT), or calcium supplementation. Non-pharmacological interventions were included if they were based around diet alteration or dietary supplements, exercise, education or consultation. Interventions were excluded if they did not directly target RED-S, if their outcome measures were not relevant to RED-S-related outcomes (see outcome eligibility criteria below), or if they focused exclusively on psychological measures (such as disordered eating, eating disorders, body image) without addressing the broader spectrum of RED-S symptoms.

#### Outcomes

Studies were included if they utilised outcomes that included measures related to menstrual resumption, energy or dietary improvement related outcome measures, physiological measures related to REDS for instance, weight, fat mass, body percentage weight, cortisol, identification of hormones, BMD. Alternative studies could be included inf they identified physical or functional measures change (e.g., strength improvements) or psychological measures (e.g., mood, body image scale).

#### Study Design

Studies were included if they reported on the experience of, or outcomes from an intervention-based study. Experimental, quasi experimental and pre-experimental designs were included. Conference proceedings, thesis and ongoing research were included to reduce the risk of publication bias. Studies were excluded if they did not report on the experience of, or outcomes from, an intervention-based study. Non-experimental designs such as case studies, observational studies, cross-sectional studies, and purely descriptive research were excluded. Reviews, editorials, opinion pieces, and commentaries were also excluded. Studies that were unpublished without accessible data, duplicate publications, and conference abstracts without full data were excluded unless further data were available.

#### Other criteria

Date restrictions were not applied to the search dates. No restriction on language was made.

### Search Strategy

A blind search by two authors (AW,AS), supported by the management software Covidence© was undertaken from a total of five electronic databases. This included; CINHAL, MEDLINE, SportDiscus, ERIC, Embase from inception until July 2025. The following key words were utilised; Relative energy deficiency in sport, OR RED-S, OR female athlete triad OR low energy deficiency OR amenorrhea OR Oligomenorrhea OR menstrual disturbance AND Intervention OR treatment OR therapy AND female OR women. In addition to this, three electronic search engines including Google Scholar, ScienceDirect and Findit.Bham were searched for the first 30 pages using the terms ‘females and relative energy deficiency and sport’. Grey literature was searched using the Grey Matters search engine. Citation chasing was undertaken with all included articles and all identified previous reviews.

### Selection process

Two blind authors (AW, AS) undertook the selection process using the Covidence© software. A separate academic with experience in systematic reviews was available to arbitrate discussions when a decision could not be made. Both authors entered decisions by title, then abstract then through reading full text independently.

### Data items

A pre-determined extraction data tool devised in Microsoft Excel identified specific variables that included demographic variables (study title, journal, design, type of intervention, geographical location, population, age, sport, athlete level, participant identifying group, if a clinical diagnosis was obtained, and method of assessment). Intervention variables were identified according to Tidier guidelines^21^, tabulated and outcome measures (identification of all outcomes, and identification of primary and secondary outcome measures) were considered which were then grouped by domains.

### Study risk of bias assessment

Study risk of bias assessment was undertaken using the ROB-2 tool^22^ for randomised control trials, or the Robins-I Version 2 tool^23^ for non-randomised control trials. For other types of design or instance case control or case series the JBI critical appraisal tools were utilised (https://jbi.global/critical-appraisal-tools). See supplementary file for the assessment.

### Effect measures

Mean differences were considered where possible for all outcome measures.

### Synthesis

A narrative synthesis documented results by outcome domain. The following outcome domain areas were identified: (a) Physiological (including, menses and menses restoration, weight, cortisol, identification of hormones, BMD). (b) Physical and functional (including measures of strengths or function) and (c) psychological outcomes (including mood, body image scale, or eating disorder questionnaire). Meta-analysis was possible and conducted for three outcome measures as part of the synthesis for non-pharmacological interventions. No other meta-analysis was possible due to the limited evidence currently available.

### Certainty assessment

GRADE^24^ as used to assess certainty of outcome measure results and supplement narrative synthesis and meta-analysis.

### Equity, diversity, and inclusion statement

Authors used the narrative synthesis to identify common demographics and intervention characteristics. The group selected within the eligibility and represented across studies are reasonably homogenous due to the individuals representing studies mainly from the USA, from University based settings and representing female endurance and distance sports/athletes.

### Patient and public involvement

No patient or public involvement in this review was undertaken.

## Results

### Search output

A total of 3156 articles were examined by both reviewers. A total of 261 were input into Covidence database of which 162 were identified for screening and 19 studies met the inclusion criteria for this systematic review. This comprised of 15 ^25–39^ (15/19, 79%) non-pharmacological interventions and 4 ^40–43^(4/19, 21%) pharmacological interventions. See Figure 1 for the PRISMA flow diagram.

**Figure 1.**
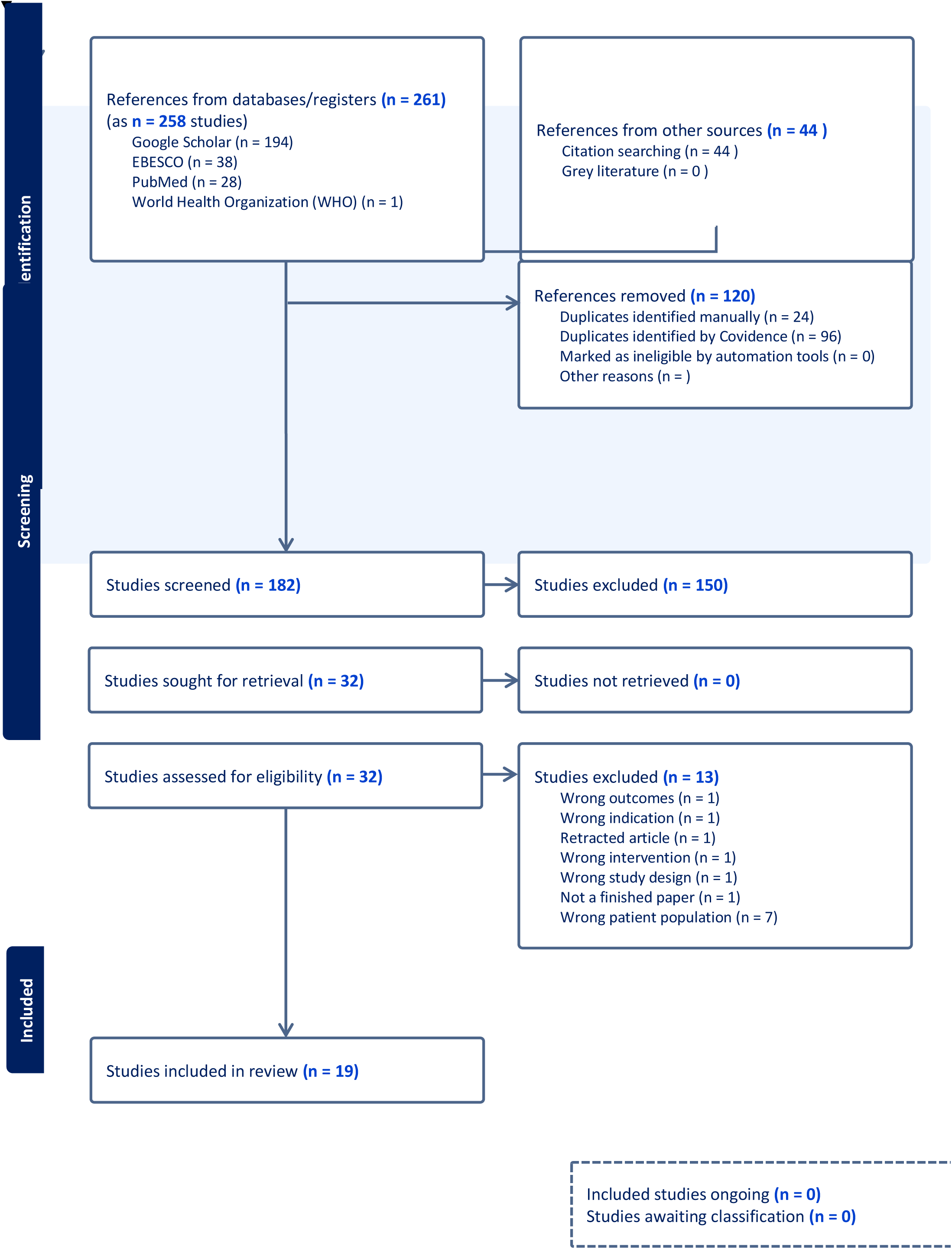
A PRISMA 2020 flow diagram produced by Covidence to identify the blind search process undertaken by two authors.

**Figure 2.**
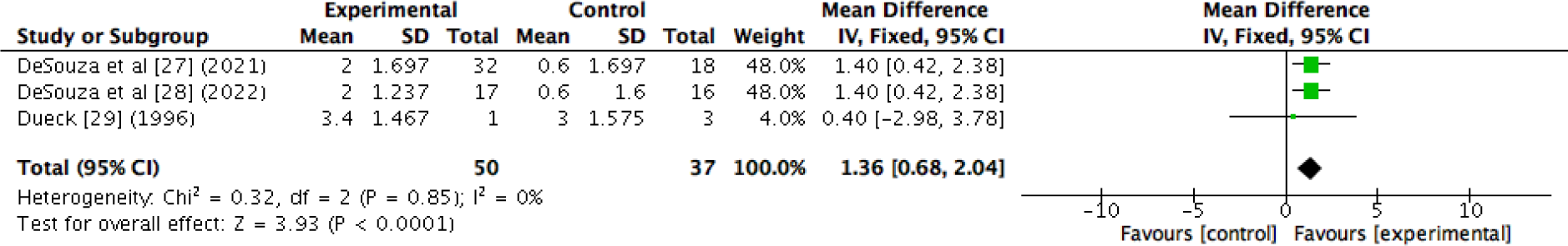
A meta-analysis showing the benefit of nutrition-based interventions on an individual’s fat mass.

**Figure 3.**
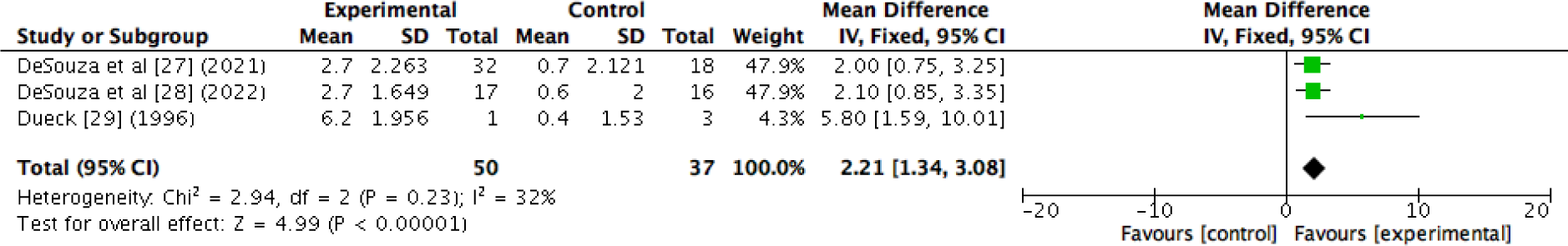
A forest plot showing the benefit of nutrition-based interventions on percentage body fat.

### Demographics

A total of 759 female participants were included (n= 759). This included individuals most often in the age bracket 18-35 years. Where possible (n=12 studies, n=473 participants) an aggregated mean age was calculated as 20.9 years. The most included group of females was female ‘athletes’ (n=12/19, 63%), followed by ‘exercising’ or ‘active’ females (n=5/19, 26%). Non-pharmacological trials included 9 (9/15, 60%) as athletes, 5 as ‘exercising’ or ‘active’ females (5/15, 33.33%) and 1 (1/15, 7%) identified without classification. Pharmacological trials included athletes (3/4, 75%) and one study included ballet dancers (1/4, 25%). The classification of included sports most often related to a multi-sport 7/19 (37%; 6/15, 40% for non-pharmacological and 1/4, 25% pharmacological) or multi-sport with the term endurance or distance 10/19 (53%; 8/15, 53% for non-pharmacological and 2/4, 50% for pharmacological). The most common specific sport mentioned was running, followed by cycling. Although exact numbers for these specific sports would be hard to determine due to the use and inclusion of multi-sports. The most common country location for studies was the USA (n=12/19, 60%) including 10/15 (67%) for non-pharmacological interventions and 2/4 (50%) for pharmacological, this was followed by Germany (n=3/19, 16%) all coming from non-pharmacological interventions. See Table 1 for a summary of non-pharmacological interventions and Table 2 for a summary of pharmacological interventions.

**Table 1.** Summary characteristics of non-pharmacological interventions.

| Authors | Study Design and classification. | Sample and sample size | Sport, athlete level and geographical location. | Participant identification (RED-S, Amennhorrea, LEA, menses, etc) | Comparator or Control | Intervention Duration (weeks) | Outcome measures | Results |
| --- | --- | --- | --- | --- | --- | --- | --- | --- |
| Arends et al <sup>25</sup> (2012) | Design: Five year retrospective cohort study. Classification: Non-Pharmacological | Female collegiate athletes (n=51) (36 oligomenorrheic, 15 amenorrheic). Age range: 18-21 years. | Sport: Multi-sport. Athlete level: Collegiate. Country: USA. | Menstrual disturbances (oligomenorrhea, amenorrhea) | Within-group comparison: athletes who did vs. did not recover menses | 260 | Primary outcome: Time to restoration of menses (ROM). Secondary outcomes: weight, BMI changes. | 17.6% (9/51) of athletes regained menses during follow-up; Mean time to ROM = 15.6±2.6 months; ROM group had significantly greater weight gain (5.3±1.1 vs 1.3±1.1kg), % weight gain (9.3±1.9% vs 2.3±1.9%), and BMI increase (1.9±0.4 vs 0.5±0.4kg/m<sup>2</sup>) compared to non-ROM athletes; Weight gain predicted ROM (OR 1.25, 95% CI 1.01–1.56). |
| Cialdella-Kam et al <sup>26</sup> (2014) | Design: Intervention study. Classification: Non-pharmacological (carbohydrate-protein supplement). | Female athletes (n=8). Age range: 19-27 years. Control: Mean 23.1 ± 4.3 years. ExMD: Mean 22.6 ± 3.3 years. | Sport: Distance runners. Athlete level: Competitive/trained. Country: USA. | Exercise-associated menstrual dysfunction (ExMD); amenorrhea | Pre-Post intervention comparison; eumenorrheic control group at baseline | 24 | Primary outcome: Resumption of menses. Secondary outcomes: reproductive & thyroid hormones; energy intake (EI); energy availability (EA); bone mineral density (BMD); bone content; muscle strength/power; protein metabolism; mood (POMS). | 7 of 8 resumed menses, <8-month ExMD showed greater skeletal improvements; Increased EA and EI (not statistically significant); Minimal bone/muscle changes; mood improved slightly; ExMD preceded >8mo had lower BMD. |
| De Souza et al <sup>27</sup> (2021) | Design: Randomised control trial. Classification: Non-Pharmacological (diet). | Exercising women (n=76). Age range: 19-23 years. Oligo/Amen + Cal: Mean 21.3±0.5 years. Oligo/Amen Control: Mean 20.7±0.5 years. | Multi-event (running, cycling, triathlon). Country: USA. | RED-S related menstrual dysfunction (amenorrhoea/oligomenorrhoea) | Standard diet vs increased energy intake | 48 | Primary outcome: Recovery of menses. Secondary outcomes: LH pulse frequency; Energy Availability; Estradiol. | Increased energy intake group had significantly higher rate of menstrual recovery (79%) and LH pulse frequency improvement than controls (29%) |
| De Souza et al <sup>28</sup> (2022) | Design: Randomised control trial. Classification: Non-pharmacological (energy intake intervention) | Exercising women (n=76) (Oligo/Amenorrhea + Cal: n=40; Control: n=36). Age range: 20-22 years. OV reference: Mean 23.4 ± 0.7 years. Oligo/Amen + Cal: Mean 21.3 ± 0.5 years. Oligo/Amen: Mean 20.7 ± 0.5 years. | Endurance activities (running, cycling, swimming), ball sports, aesthetic sports, power-sport. Country: USA. | Oligo-amenorrhea; risk of RED-S | Control group without intervention | 52 | Primary outcome: Bone mBMD. Secondary outcomes: Body mass/BMI/fat mass/fat %/IGF-1; menstrual frequency. | Intervention increased weight, BMI, fat mass, fat %, IGF-1, and menstrual frequency (P<0.05) vs control; Total body & spine aBMD: no change vs control; Femoral neck and hip aBMD decreased over 12 mo in both groups |
| Dueck et al., <sup>29</sup> (1996) | Design: Intervention study with matched controls. Classification: Non-pharmacological | Female athletes (n=4) (1 amenorrheic; 3 eumenorrheic comparators). Age range 19-20 years. | Sport: Endurance running. Athlete level: Collegiate. Country: USA. | Amenorrhea | Eumenorrheic controls | 15 | Primary outcome: Energy balance. Secondary outcomes: Body fat percentage, fasting LH & cortisol, menstrual function | Amenorrheic athlete regained normal hormonal levels and menstrual cyclicity with improved energy balance, increased fat mass, increased LH, and reduced cortisol. Control athletes showed stable hormones/fat. |
| Fahrenholtz et al <sup>30</sup> (2023) | Design: Non-randomised controlled trial. Classification: Non-pharmacological (lectures & counselling). | Female athletes (n=50). Age range: 18-35 years. FUEL: Mean 24.1 ± 4.7 years. CONTROL: Mean 25.3 ± 4.8 years. (32 in FUEL intervention; 18 in control group) | Sport: Endurance sports (long-distance running, orienteering, triathlon, cycling, cross-country skiing, biathlon). Athlete level: Recreational and competitive. Country: Norway, Sweden, Ireland, Germany. | Risk of RED-S (LEAF-Q ≥8) | Control group without lectures/counselling | 16 | Primary outcome: Sports nutrition knowledge (interviews & self-perceived). Secondary outcomes: Dietary behaviour (7-day weighed food record, habit questions). | Strong evidence of improved nutrition knowledge (28% increase); weak evidence of improved behaviour (e.g., carbohydrate intake days/week improved marginally) |
| Fredericson et al.<sup>31</sup> (2023) | Design: Intervention study. Classification: Non-Pharmacological (nutrition education). | Collegiate distance runners (n = 56 retrospective phase; n=78 intervention phase) | Sport: Distance running (≥800m). Athlete level: Collegiate. Country: USA. | At high-risk of Bone Stress Injury (BSI) | compare bone stress injury rates in historical and intervention phases | 208 | Primary outcome: Incidence of BSI. Secondary outcomes: Energy intake, bone density. | The intervention resulted in a significant reduction in BSI alongside improved energy intake and bone mineral density |
| Guebels et al.<sup>32</sup> (2014) | Design: Intervention with control group. Classification: Non-Pharmacological (dietary supplement). | Active women (n=17) (8 ExMD; plus 9 eumenorrheic active controls). Age range 19-29 years. | Sport: Endurance sports. Athlete level: Endurance trained (≥7h/week). Country: USA. | ExMD; eumenorrhea | eumenorrheic control at baseline | 24 | Primary outcome: Resting metabolic rate (RMR). Secondary outcome: Energy intake (EI); total energy expenditure (TEE); energy availability (EA); energy balance (EB); weight | ExMD had higher baseline TEE and more negative EB; RMR higher in ExMD; increased weight; No significant changes in EA, EB, or RMR post-intervention; EA ranged 28.2–36.7 to 30–45.4 kcal/kgFFM/d. |
| Kopp-Woodroffe et al.<sup>33</sup> (1999) | Design: Intervention pilot study. Classification: Non-pharmacological (diet and exercise) | Active females (n=4). Age range: 19-34 years. | Sport: Running, swimming, cycling, weight-training. Athlete level: Competitive (training 7+hr/week). Country: USA. | athletic amenorrhea | Pre-Post intervention comparison | 20 | Primary outcome: Energy balance. Secondary outcomes: EI, EE, serum biomarkers | Improved energy balance; increase body weight; increased body fat; improved nutrient status; resumption of menstrual function in 3 participants |
| Lagowska et al.<sup>34</sup> (2014) | Design: Non-randomised intervention. Classification: Non-pharmacological. | Female athletes (n=52) (31 athletes, 21 ballet dancers). Age range: 16-21 years. Ballet: Mean 17.1±0.9 years. Athletes: 18.1±2.6 years. | Sport: Ballet & endurance sport (triathlon, rowing, swimming). Athlete level: Competitive/trained. Country: Poland. | Menstrual disorders (amenorrhea/oligomenorrhea) | Baseline vs post-intervention; dancers vs athletes within-study | 36 | Primary outcome: Restoration of menses. Secondary outcomes: Energy & nutrient intake; TEE; EA; body composition; hormones (LH, FSH, E2, P, TSH, PRL, SHBG, leptin); resting metabolic rate (RMR) | Regular menses resumed in 10/52 participants; EA & energy intake increased; RMR low in dancers; Weight/BMI/body composition improved in dancers; not athletes; Higher body fat linked to menstrual recovery. |
| Lagowska et al.<sup>35</sup> (2014) | Design: Non-randomised intervention. Classification: Non-pharmacological. | Female athletes (n=31). Age range: 15-21 years. Mean: 18.1±2.6 years. | Sport: Rowing, synchronised swimming, and triathlon. Athlete level: Professional. Country: Poland. | Amenorrhea; oligomenorrhea (low energy availability related) | Pre/post within-subject; no independent control group | 12 | Primary outcome: Energy intake. Secondary outcomes: TEE, EA, EB, body composition, LH, FSH, oestradiol, progesterone | Significant improvement in energy intake, EA, EB, and LH hormone profiles, but no restoration of menstrual cycles within 3 months |
| Mallinson et al.<sup>36</sup> (2013) | Design: Case report. Classification: Non-Pharmacological (diet). | Exercising women (n=2). Age range 19-24 years. | Sport: Multi-event (running, weight-training, rock climbing, hiking, downhill skiing, dancing). Athlete level: Recreational (exercise 7+ hr/week). Country: USA. | Functional Hypothalamic Amenorrhea (FHA) | Pre-post case report | 6.9 | Primary outcome: Recovery of menses. Secondary outcomes: Energy intake; total body mass. | P1: +4.2 kg body mass (8%), P2: +2.8 kg (5%); both showed ↑ REE, ↑ T3 and leptin, ↓ ghrelin; menstrual resumption at day 23 (P1) and 74 (P2); ovulation and cycle regularity returned closely with weight gain |
| Meyer et al.<sup>37</sup> (2025) | Design: Clinical practice report. Classification: Non-Pharmacological (consultation programme). | Female Athletes (n=58). Age range: 13-24 years. Mean: 18.3 ± 5.5 years. | Sport: Multi-event (game sports, endurance, strength, martial arts, aesthetic, technical). Athlete level: Competitive (3-27 hrs/week). Country: Germany. | Suspected RED-S | Pre-post intervention comparison | 188 | Primary outcome: Improvement in RED-S symptoms. Secondary outcome: Energy intake; bone density. | Multidisciplinary approach resulted in improvement in RED-S symptoms and health outcomes |
| Michopoulos et al.<sup>38</sup> (2013) | Design: Randomised control trial. Classification: Non-pharmacological (cognitive behavioural therapy). | Women (n=17). Age range: 23-27 years. Observation: Mean 25.1 ± 1.8 years. CBT: 25.3 ± 0.6 years. | Sport: Not identified. Athlete level: Not identified. Country: USA. | Functional Hypothalamic Amenorrhea (FHA) | Observation (no CBT) | 20 | Primary outcome: Cortisol. Secondary outcomes: Leptin, TSH, total/free T3 & T4 concentrations | CBT significantly lowered cortisol (p = 0.006) and increased leptin & TSH; observation group showed no changes |
| Solstad et al.<sup>39</sup> (2025) | Design: Mixed methods; quasi-experimental. Classification: Non-pharmacological. | Female athletes (n=44). Age range 18-35 years. Evaluation questionnaire: Mean 24.6 ± 4.8 years. Qualitative interview: Mean 25.1 ± 5.8 years. | Sport: Endurance (long-distance running, orienteering, cycling, triathlon, cross-country skiing, biathlon). Athlete level: Competitive (training 20+ session/month). Country: Multicentric (Germany, Ireland, Norway, Sweden) | Relative Energy Deficiency in Sport (RED-S; at risk). Self-reported diagnosis/no clinical diagnosis. | Digital lectures & consultation vs digital lectures only | 16 | Primary outcome: Overall satisfaction. Secondary outcome: Energy, confidence, body satisfaction, knowledge | Participants reported high satisfaction and intention to recommend; no significant LEAF-Q group differences post-intervention, but improvements in menstrual and GI symptoms seen at 6 and 12 months follow-up |

Note: Body Mass Index (BMI), Lutenizing Hormone (LH), Follicle Stimulating Hormone (FSH), Thyroid Stimulating Hormone (TSH), Inulin-like Growth Factor 1 (IGF 1), Prolactin (PRL), Sex Hormone Binding Globulin (SHBG), Triiodothyronine (T3).

**Table 2.**
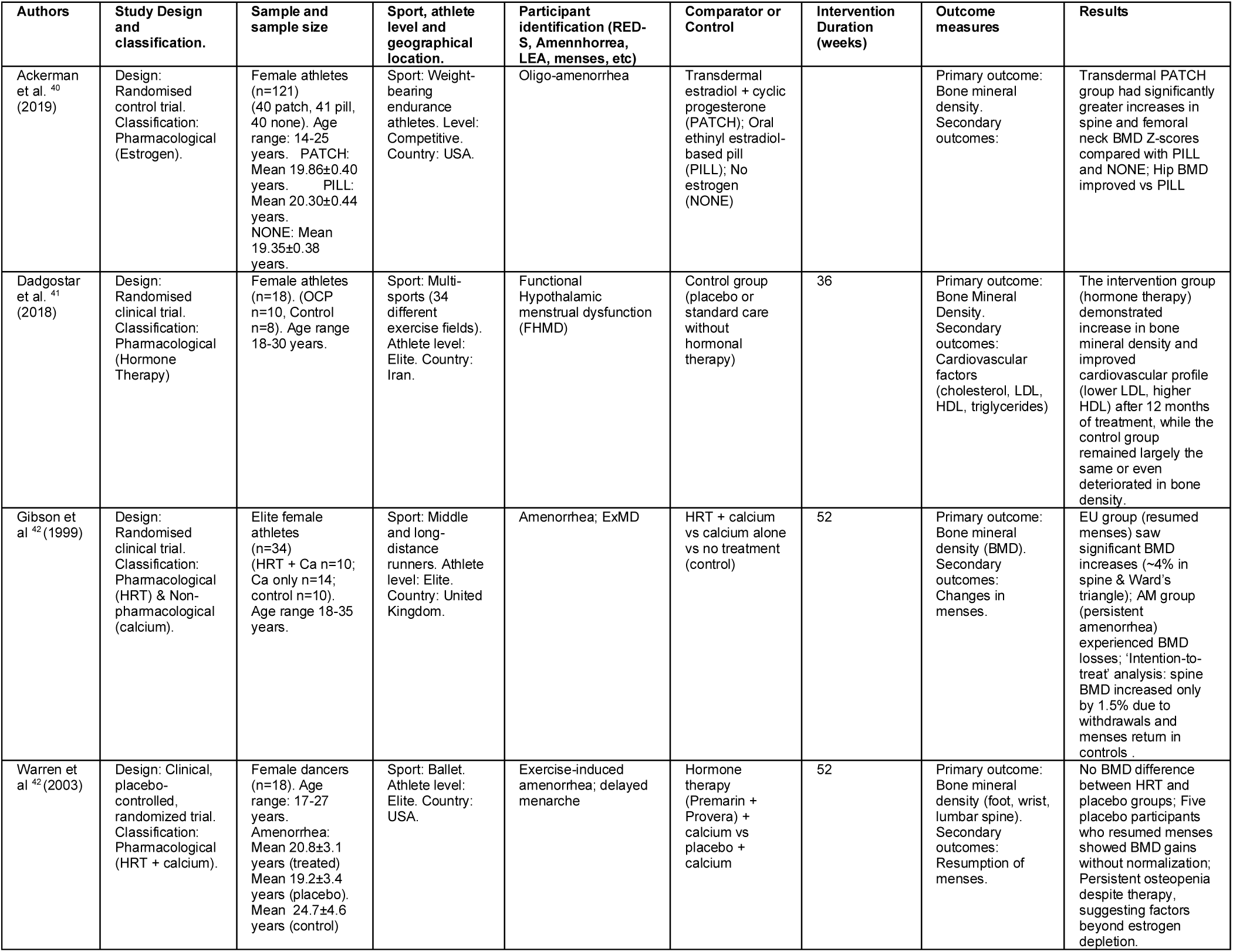
Summary characteristics of pharmacological interventions.

| Authors | Study Design and classification. | Sample and sample size | Sport, athlete level and geographical location. | Participant identification (RED-S, Amenorrhea, LEA, menses, etc) | Comparator or Control | Intervention Duration (weeks) | Outcome measures | Results |
| --- | --- | --- | --- | --- | --- | --- | --- | --- |
| Ackerman et al. <sup>40</sup> (2019) | Design: Randomised control trial. Classification: Pharmacological (Estrogen). | Female athletes (n=121) (40 patch, 41 pill, 40 none). Age range: 14-25 years. PATCH: Mean 19.86±0.40 years. PILL: Mean 20.30±0.44 years. NONE: Mean 19.35±0.38 years. | Sport: Weight-bearing endurance athletes. Level: Competitive. Country: USA. | Oligo-amenorrhea | Transdermal estradiol + cyclic progesterone (PATCH); Oral ethinyl estradiol-based pill (PILL); No estrogen (NONE) |  | Primary outcome: Bone mineral density. Secondary outcomes: | Transdermal PATCH group had significantly greater increases in spine and femoral neck BMD Z-scores compared with PILL and NONE; Hip BMD improved vs PILL |
| Dadgostar et al. <sup>41</sup> (2018) | Design: Randomised clinical trial. Classification: Pharmacological (Hormone Therapy) | Female athletes (n=18). (OCP n=10, Control n=8). Age range 18-30 years. | Sport: Multi-sports (34 different exercise fields). Athlete level: Elite. Country: Iran. | Functional Hypothalamic menstrual dysfunction (FHMD) | Control group (placebo or standard care without hormonal therapy) | 36 | Primary outcome: Bone Mineral Density. Secondary outcomes: Cardiovascular factors (cholesterol, LDL, HDL, triglycerides) | The intervention group (hormone therapy) demonstrated increase in bone mineral density and improved cardiovascular profile (lower LDL, higher HDL) after 12 months of treatment, while the control group remained largely the same or even deteriorated in bone density. |
| Gibson et al. <sup>42</sup> (1999) | Design: Randomised clinical trial. Classification: Pharmacological (HRT) & Non-pharmacological (calcium). | Elite female athletes (n=34) (HRT + Ca n=10; Ca only n=14; control n=10). Age range 18-35 years. | Sport: Middle and long-distance runners. Athlete level: Elite. Country: United Kingdom. | Amenorrhea; ExMD | HRT + calcium vs calcium alone vs no treatment (control) | 52 | Primary outcome: Bone mineral density (BMD). Secondary outcomes: Changes in menses. | EU group (resumed menses) saw significant BMD increases (~4% in spine & Ward's triangle); AM group (persistent amenorrhea) experienced BMD losses; 'Intention-to-treat' analysis: spine BMD increased only by 1.5% due to withdrawals and menses return in controls. |
| Warren et al. <sup>42</sup> (2003) | Design: Clinical, placebo-controlled, randomized trial. Classification: Pharmacological (HRT + calcium). | Female dancers (n=18). Age range: 17-27 years. Amenorrhea: Mean 20.8±3.1 years (treated) Mean 19.2±3.4 years (placebo). Mean 24.7±4.6 years (control) | Sport: Ballet. Athlete level: Elite. Country: USA. | Exercise-induced amenorrhea; delayed menarche | Hormone therapy (Premarin + Provera) + calcium vs placebo + calcium | 52 | Primary outcome: Bone mineral density (foot, wrist, lumbar spine). Secondary outcomes: Resumption of menses. | No BMD difference between HRT and placebo groups; Five placebo participants who resumed menses showed BMD gains without normalization; Persistent osteopenia despite therapy, suggesting factors beyond estrogen depletion. |

### Intervention characteristics

The most common characteristics across studies was as follows. The classification system used to identify athletes most often identified Amenorrhea (n=6/19) as a clinical inclusion criterion for participation. Common intervention components included (a) increasing energy intake/diet supplementation (n=10/19; 53%; 10/15, 67% for non-pharmacological interventions) and (b) dietary/nutritional counselling or advice (out of all studies, n=6/19, 32%) and 6/15 (40%) for non-pharmacological interventions was the most prevalent intervention component. The average duration of intervention for non-pharmacological interventions was 28.3 ± 15.8 weeks (n=12/15, 80%). The setting selected for the intervention was most often described as a ‘university’ setting (n=8/19, 42%), this included 8/15 (53%) for non-pharmacological studies. Staff involved in the interventions were most often identified as dietitians (n=12/19) 12/15 (80%) for non-pharmacological studies and 1/4 (25%) for pharmacological interventions, or as researchers (n=9/19, 47%), including 8/15 (53%) for non-pharmacological interventions and 1/4 (25%) of pharmacological evidence.

**Table 3.** A summary table for non-pharmacological studies (using Tidier sub-headings)

| Authors | RED-S diagnosis criteria used | Control or Comparator | Intervention strategy | What was provided (materials/information) | Intervention setting (where) | Duration and Dosage | Who provided (MDT involvement) |
| --- | --- | --- | --- | --- | --- | --- | --- |
| Arends et al.<sup>25</sup> (2012) | measuring estradiol (pg/ml), LH (mIU/ml), follicle-stimulating hormone (FSH; mIU/ml), thyroid-stimulating hormone (mIU/L), and prolactin (ng/ml); a urine pregnancy test | Within-sample comparison: "resumers" vs "non-resumers" following nonpharmacologic intervention | increased dietary intake and/or reduced exercise expenditure | Individualized nutrition and training recommendations to address low energy availability | university student health center | Mean time to restoration of menses: 15.6 ± 2.6 months; intervention varied per athlete | College health center providers |
| Cialdella-Kam et al.<sup>26</sup> (2014) | Exercise-associated menstrual dysfunction: ≥3 months of no menses or ≤6 cycles/year; identified as low EA from energy intake/expenditure logs | Eumenorrheic athlete controls assessed at baseline only | Daily carbohydrate-protein supplement (360 kcal/day: 54 g CHO + 20 g PRO) for 6 months | 360 kcal CHO-PRO drink; baseline and ongoing 7-day diet and activity logs to track energy balance | University-based lab/clinical setting | 6 months; supplement consumed daily; mid-intervention check at 3 months | Nutrition and exercise science research team |
| De Souza et al.<sup>27</sup> (2021) | Self-reported amenorrhea (>3 months) or oligomenorrhea (<7 cycles in 12 months or cycles of 36–89 days), with suppressed reproductive hormones; exercise ≥2 hours/week | 40 in + Cal intervention; 36 in Control - Maintenance of usual energy intake (Control arm) | Increase energy intake by 20–40% above baseline needs + nutritionist guidance and supplements | Nutrition counselling; energy bars (220–300 kcal) and pre-measured nut servings; RMR/energy needs assessments; urinary hormone logs | University-based clinical research Centre's | 12 months | Registered clinical dietitians and clinical psychologists; research team included sports endocrinology/investigators |
| De Souza et al.<sup>28</sup> (2022) | self-reported cycles; secondary FHA due to energy deficiency. Confirmed via menstrual history and hormonal measures resembling prior REFUEL paper | Oligo/Amen control group maintaining usual diet/energy intake | increased energy intake by 20–40% over baseline needs (~+352 kcal/d) | Personalized nutrition counseling plus supplemental energy sources to raise intake | Clinical nutrition research | 12 months continuous intervention | Research nutritionists & investigators |
| Dueck et al.,<sup>29</sup> (1996) | estradiol, progesterone, luteinizing hormone (LH), follicle-stimulating hormone (FSH), and cortisol | 3 matched eumenorrheic endurance athletes over same 15-week period | reduced training load + nutritional supplementation | 360 kcal/day sport nutrition beverage; 1 additional rest day per week | University research facility | 15 weeks; daily supplement; 1 added rest day/week | exercise physiologists and nutrition scientists |
| Fahrenholtz et al.<sup>30</sup> (2023) | LEAF-Q ≥ 8 (low energy availability); EDE-Q < 2.5 (low risk of eating disorders) | control group (n = 18), same assessments without lectures or counseling | Combination of weekly online sports nutrition lectures (16 total) and biweekly individual athlete-centered nutrition counseling | Lecture materials (evidence-based content), telephone/online interviews, personalized nutrition counseling every 2 weeks | Digital delivery across Norway, Sweden, Ireland, and Germany | 16 weeks total; 16 lectures and 8 counseling sessions across that period | Delivered by researchers and practicing sports nutritionists |
| Fredericson et al.<sup>31</sup> (2023) | Female Athlete Triad risk screening to identify those at risk for low EA and BSI | Historical control: retrospective data from 2010–2013; Intervention group followed prospectively 2016–2020 | Nutrition education sessions targeting energy availability and bone-building nutrients; individualized follow-up for high-risk athletes | Team-based presentations on fueling and bone health; individual sessions for Triad-risk athletes | Two NCAA Division I collegiate running programs—team and individual sessions on site | 7 years overall: pilot 2013–2016; intervention implemented 2016–2020 | Sports dietitians and sports medicine/research personnel led education; team coaches and athletic trainers involved |
| Guebels et al.<sup>32</sup> (2014) | menstrual status was confirmed by presence/absence of ovulation; amenorrheic (no menses > 90 days), oligomenorrheic (cycle intervals > 35d) or eumenorrheic (10–13 cycles/year or intervals of ~28 days). | Active eumenorrheic women (n = 9) served as controls | +360 kcal/day supplement for 6 months to restore energy balance | Daily 360 kcal supplement; 7-day diet and exercise logs to monitor intake and expenditure | University lab-based research setting | 6 months; supplement consumed daily; RMR assessed at baseline and 6 months | exercise physiologists/nutrition researchers |
| Kopp-Woodroffe et al.<sup>33</sup> (1999) | Self-report menstrual history; reproductive and thyroid hormone analysis (serum progesterone, serum estradiol) | Pre-Post intervention | Decrease EE (1 rest day/week) + Increase EI (11 oz CHO/Pro/Fat + micronutrient supplement per day) | Dietary supplement; calibrated diet scale; measuring cups | Home environment & lab | 20 weeks | Researchers |
| Lagowska et al.<sup>34</sup> (2014) | Secondary amenorrhea (>6 months) or oligomenorrhea (>35-day cycles), low EA based on diet and exercise logs | Pre-post intervention comparison | individualized diet plan to increase energy and nutrient intake without altering training | Monthly dietitian consultations; tailored diet based on energy/nutrient needs; 7-day dietary and exercise records | University nutrition lab | 9 months, with evaluations at 3 (same population not included in review analysis), 6, and 9 months | Dietitians and research team (nutrition and exercise specialists) |
| Lagowska et al.<sup>35</sup> (2014) | Amenorrhea or oligomenorrhea (NH subtyping); lab exclusion of prolactin, thyroid, ovarian, androgen, hCG abnormalities | Pre-post intervention comparison | 3-month individualized dietary intervention to increase energy and nutrient intake, without reducing training | Tailored diet plans increasing energy (~+234 kcal), protein (+8 g), carbs (+66.8 g), calcium, vit A/D/C/folate; 7-day dietary/exercise logs | University nutrition lab | 3 months; moderate daily energy/nutrient increases | Dietitians and exercise science research team |
| Mallinson et al.<sup>36</sup> (2013) | Amenorrhea ≥3 months; daily urinary excretion of E1G and PdG metabolites for 28 day period. | Pre-post case report | Increased dietary intake 20–30% above baseline TEE | Personalized caloric increase via energy bars (250–300 kcal): +276 kcal/day for P1; +1,881 kcal/day for P2; ongoing dietary monitoring and support | University laboratory / clinical research facility | Until menses resumed: 23 days for P1; 74 days for P2 | Women's Health & Exercise Lab research team; exercise physiologists and nutrition researchers |
| Meyer et al.<sup>37</sup> (2025) | RED-S suspicion via CAT1 (BMI, body fat %, weight loss, menstrual disorders, bone health history) | Pre-post comparison | Interdisciplinary outpatient consultation including sports medicine, gynecology, psychosomatic medicine, dietetics, and psychiatry | Medical records review, anthropometry, performance diagnostics, lab tests, dietary records, psychological and gynecological assessments; tailored treatment/referral pathway | University Hospital Tübingen outpatient clinics (sports medicine, gynecology, psychosomatic, psychiatry, dietetics) | Mean treatment duration ~15 months (up to 36); initial high intensity in 2–3 months, follow-ups 1–2 years; appointments ranged from 2–15 in sports med, up to 4 in psychiatry | sports medicine physicians, gynecologists, psychosomatic/child & adolescent psychiatrists, registered dietitians, and health educators |
| Michopoulos et al.<sup>38</sup> (2013) | FHA defined by >3 months of amenorrhea; exclusion of other causes via serum E2 and FSH; prolactin, testosterone, DHEA, TSH, and urinary hCG | Randomized to CBT intervention (n=8) or observation control (n=9) | weekly cognitive-behavioral therapy sessions | Structured CBT program focused on stress, dieting/exercise beliefs, and lifestyle behaviors | Clinical research center at Emory University | Approximate 20 weekly sessions (~5 months) (number inferred from context) | clinical psychologists specializing in CBT under research supervision |
| Solstad et al.<sup>39</sup> (2025) | LEAF-Q score ≥ 8 (risk of RED-S); EDE-Q < 2.5 (low risk of eating disorder) | 33 participants partook in digital lectures & consultation, 11 participants completed digital lectures only | digital sports nutrition lectures & individual consultations | Weekly online lectures, bi-weekly individualised nutrition counselling via motivational interviewing, focussed on RED-S, nutrition intake, and menstrual health. | Home environment - online lectures; zoom video | 16 weeks | Sports nutritionists |

**Table 4.**
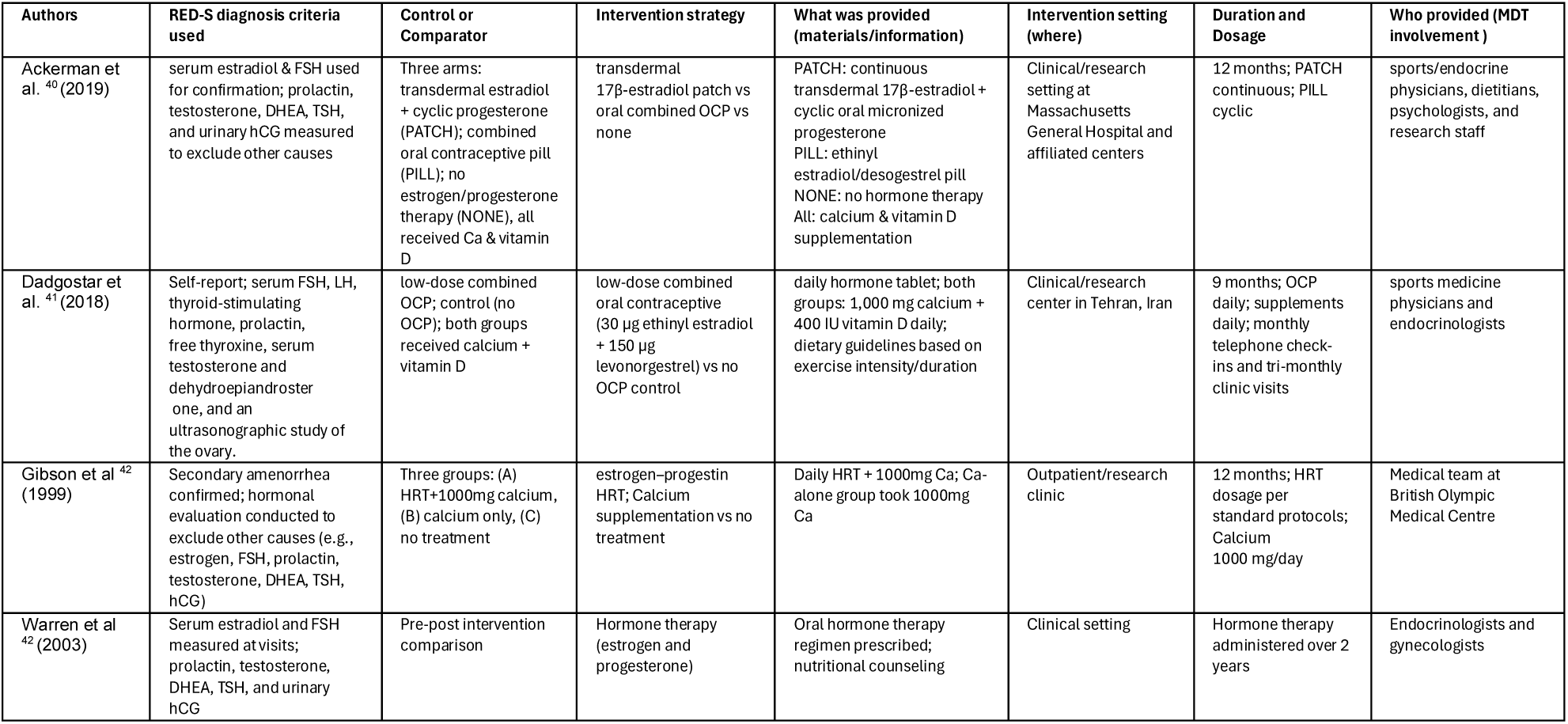
Tider summary table for intervention studies.

### Synthesis & Certainty Assessment

Findings are provided below for non-pharmacological evidence and pharmacological evidence. A supplementary file is available that provides the assessment of critical appraisal (quality) and certainty assessment.

### Non-pharmacological

Among the studies assessing non-pharmacological interventions, the most frequently reported outcomes were menstrual function recovery (n= 8/15, 53%), improvements in energy availability (EA) (n= 5/15, 33%), changes in body composition (n= 7/15, 47%), and alterations in relevant biomarkers (n= 9/15, 60%). These were reported below.

### Menstrual function recovery

Menstrual function recovery was utilised as an outcome measure in eight (n= 8/15, 53%) of the included papers. Seven (n= 7/8, 88%) reported statistically significant improvements (p-values ranging from <0.01 to 0.05), with large, estimated effect sizes (0.8–1.2). One case report lacked statistical analysis but described clinically meaningful recovery in both participants. All nine studies were judged to show clinically meaningful outcomes, primarily associated with increased EA through nutritional (n= 5/8, 63%) or non-pharmacological interventions (n= 1/8, 13%) or both nutritional and non-pharmacological interventions (n= 2/8, 25%). Six (6/8, 75%) studies reported number of intervention participants that experienced improved or recovery of menses. Of the 142 intervention group participants, 47 (47/142, 33%) had partial or full recovery of menses.

**<u>Confidence in evidence:</u>**

Summary of evidence contributing to certainty rating: GRADE study ratings included 1 very low, 5 low, and 2 high.

Overall confidence rating: Low, although findings can be taken with reasonable confidence due to consistency of evidence.

**Energy availability:**

Changes in EA were evaluated in five studies (n= 5/15, 33%). All five reported statistically significant improvements (p-values ranging from <0.01 to 0.05), with estimated effect sizes between 0.5 and 1.0. Each study demonstrated clinically meaningful improvements, achieved through increased caloric intake (n=5/15, 33%).

**<u>Confidence in evidence:</u>**

Summary of evidence contributing to certainty rating: GRADE study ratings included 5 low.

Overall confidence rating: Low, although findings can be taken with reasonable confidence due to consistency of evidence.

**Body composition measures:**

Body Composition was measured in seven (n= 7/15, 47%) of the non-pharmacological papers. Six (n= 6/7, 86%) reported statistically significant improvements in at least one parameter—such as body weight, fat mass, or body fat percentage—with p-values ranging from <0.001 to 0.05 and effect sizes between 0.5 and 2.5. One study did not report statistical significance but showed large, estimated effects. All seven studies were considered clinically meaningful, with improvements primarily associated with increased energy intake. Meta-analysis was undertaken for the measures of changes in fat mass and percentage of body fat.

**<u>Confidence in evidence:</u>**

Summary of evidence contributing to certainty rating: GRADE study ratings included 5 low and 2 very high.

Overall confidence rating: Moderate, although findings can be taken with reasonable confidence as evidence is supported by meta-analysis.

**Hormonal and blood biomarkers:**

Biomarkers related to RED-S were analysed in nine (n= 9/15, 60%) studies. Eight (n= 8/9, 89%) reported statistically significant improvements (*p*< 0.05), with effect sizes ranging from 0.5 to 0.8. The most assessed markers included serum leptin, triiodothyronine (TT3), cortisol, oestradiol, and luteinizing hormone (LH) pulsatility. One study, while lacking statistical testing, demonstrated substantial changes in multiple biomarkers and was considered clinically meaningful. All nine studies reported clinically meaningful outcomes, supporting the utility of biomarker monitoring in evaluating recovery in RED-S and the effectiveness of dietary and non-pharmacological interventions. A meta-analysis (see Figure 4) was possible regarding the reporting of TT3 by three studies.

**Figure 4.**
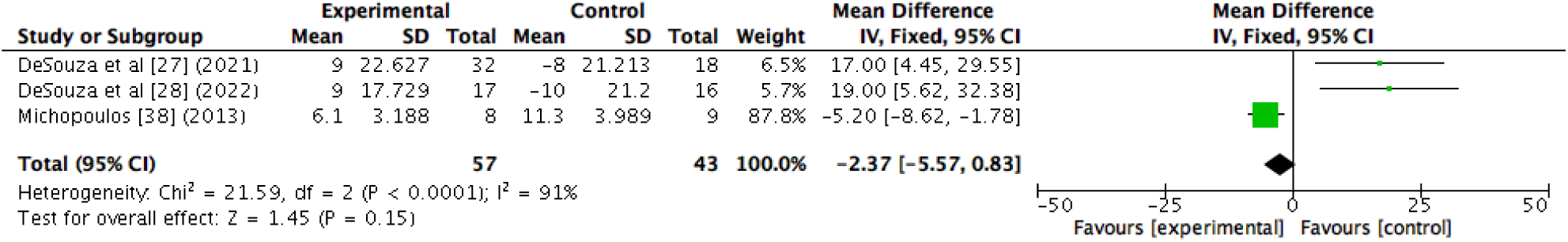
A forest plot showing the T3 biomarker changes from non-pharmacological interventions.

**<u>Confidence in evidence:</u>**

Summary of evidence contributing to certainty rating: GRADE study ratings included 6 low, 3 very high and 8 with moderate effect.

Overall confidence rating: Low, evidence effected by quality downgrades and meta-analysis favouring control group. Further evidence is required.

**Pharmacological:**

Studies examining pharmacological treatments commonly assessed BMD (n= 4/4, 100%), hormonal profiles (e.g., oestrogen, L, follicle-stimulating hormone (FSH)) (n= 3/4, 75%), recovery of menstrual function (n= 3/4, 75%) and bone turnover markers (n= 1/4, 25%) as indicators of skeletal health and remodelling.

**Bone mineral density markers:**

BMD was assessed in four (n= 4/4, 100%) pharmacological studies, with two (n= 2/4, 50%) reporting statistically significant improvements (*p*=0.021 and 0.05) and moderate effect sizes of approximately 0.8. Both significant studies demonstrated clinically meaningful increases in BMD, primarily following oestrogen replacement therapy. However, two studies did not find significant changes in BMD.

**<u>Confidence in evidence:</u>**

Summary of evidence contributing to certainty rating: GRADE study ratings included 1 very low, 1 low, 1 high, and 1 very high.

Overall confidence rating: Moderate, findings can be taken with reasonable confidence due to consistency of evidence.

**Hormonal profiling:**

Hormonal profiles were evaluated in four (n= 4/4, 100%) studies, with two (n= 2/4, 50%) showing significant improvements (*p*< 0.05) in key hormones such as oestradiol and progesterone, with effect sizes around 0.7–0.8. One study reported normalization of hormonal levels in treatment groups despite no baseline significance testing. Three (n= 3/4, 75%) studies assessing hormonal outcomes were considered clinically meaningful.

**<u>Confidence in evidence:</u>**

Summary of evidence contributing to certainty rating: GRADE study ratings included 1 very low, 1 low, and 1 very high, 1.

Overall confidence rating: Moderate, findings can be taken with reasonable confidence due to consistency of evidence despite quality.

**Menstrual function recovery:**

Menstrual function recovery was reported in three (n= 3/4, 75%) studies, all demonstrating statistically significant improvements (p-values ranging from *p*<0.05 to 0.0001). These improvements were deemed clinically meaningful, indicating effective restoration of menstrual function with pharmacological treatment.

**<u>Confidence in evidence:</u>**

Summary of evidence contributing to certainty rating: GRADE study ratings included 1 very low, 1 low, 1 high, 1 with large effect and dose-response.

Overall confidence rating: Moderate, findings can be taken with reasonable confidence due to consistency of evidence.

**Bone turnover markers:**

Bone turnover markers were evaluated in one (n= 1/4, 25%) study, which found significant changes in markers including P1NP and IGF-1 (*p*= 0.016), with clinically meaningful effects observed.

**<u>Confidence in evidence:</u>**

Summary of evidence contributing to certainty rating: GRADE study ratings included 1 study rated as high certainty.

Overall confidence rating: Low, further evidence is required to repeat results.

## Discussion

This review provides initial insight and certainty of evidence considering the effectiveness of non-pharmacological interventions compared to pharmacological interventions. Non-pharmacological interventions demonstrated consistent improvements across multiple outcome domains, including recovery of menstrual function, EA, body composition, and hormonal biomarkers. Pharmacological interventions, utilising oestrogen therapy and hormone replacement showed some effectiveness in improving BMD, restoring menstrual function and enhancing hormonal profiles. However, the certainty of evidence across both intervention types was limited by methodological weakness and number of contributing studies. The discussion now provides consideration to the main findings by intervention type.

### Non-pharmacological interventions

#### Menstrual function recovery

Despite the overall low quality of evidence, all studies consistently demonstrated clinically meaningful improvements in menstrual function following non-pharmacological interventions. These findings align with broader physiological and clinical literature. Mechanistically, energy deficiency resulting from insufficient fuel availability suppresses the hypothalamic-pituitary-ovarian (HPO) axis, leading to menstrual dysfunction^44^ . Supporting this, research shows that LH pulsatility is disrupted when EA drops below a clinical threshold, highlighting that EA rather than body fat or exercise alone is the primary regulator of reproductive function in active women^45^ . Clinical evidence further supports this, with several studies showing that dietary or training modifications can restore menses within several months in affected athletes ^46,47^.

Current studies vary widely in how recovery is defined, often relying on menstrual bleeding alone^46–49^. which does not confirm ovulation or hormonal restoration^50,51^. Menstrual bleeding alone does not guarantee hormonal balance or ovulation. Highlighting the need for standardised, hormonally validated definitions in future research. Future research should adopt standardised, hormonally validated definitions of menstrual recovery to improve consistency and clinical relevance. In addition, longer follow-up periods and consistent use of hormonal markers are needed to assess the long-term health impacts of interventions on reproductive, bone, and overall health in female athletes with RED-S.

#### Energy availability

EA was identified as improving following interventions. These findings are strongly supported by past research demonstrating the central role of EA in hormonal, reproductive, and metabolic regulation. Low EA has been shown to impair LH pulsatility, suppress resting metabolic rate, and reduce oestrogen and IGF-1 levels, negatively affecting reproductive and bone health^52,53^. Importantly, even modest increases in EA can reverse these changes, restoring LH pulsatility, resuming menstrual function, and normalising metabolic function, highlighting EA as a key modifiable factor in both the development and recovery from RED-S. Improvements in bone health have also been observed with prolonged energy restoration, though severe cases may require combined nutritional and pharmacological intervention^54^ .

Accurate measurement of EA remains a major challenge due to the difficulty of precisely assessing dietary intake, exercise, and fat-free mass. Common reliance on self-reported data and indirect proxy markers, such as menstrual dysfunction, resting metabolic rate, and hormonal changes (reduced leptin and T3), introduces variability and error, limiting confidence in current findings^17,52^. Therefore, future research needs improved, objective methods to accurately quantify EA in free-living athletes, alongside standardized and sensitive biomarkers, to strengthen the evidence base and guide effective RED-S management.

#### Body composition

Consistent and clinically relevant improvements in body composition were observed across studies. Past evidence highlights the complexity of using body composition as a primary indicator of recovery in RED-S. Although improvements in fat mass and body weight may reflect enhanced EA and nutritional rehabilitation^17^, they are not universally required for physiological recovery. Critically, key outcomes such as the return of menses and improvements in BMD, central to RED-S recovery, can occur independently of significant changes in body composition^55^. This is particularly relevant in athletes who are constitutionally lean, where minimal or no changes in fat mass may accompany full recovery of hormonal and metabolic function^56^. Moreover, reliance on body composition alone may overlook meaningful clinical progress or delay appropriate intervention if weight change is minimal. As such, current consensus, including the 2023 IOC statement on RED-S^1^ emphasises that body composition should not be used in isolation to define recovery. Instead, it should be interpreted within a broader clinical framework, incorporating menstrual status, hormonal markers, non-pharmacological changes, and performance indicators. This multidimensional approach is essential for accurately monitoring recovery and tailoring individualised treatment strategies in athletes affected by RED-S.

#### Biomarkers

Endocrine markers, particularly serum leptin and triiodothyronine (TT3), along with LH and oestradiol, showed consistent, clinically meaningful improvements across studies, indicating recovery of metabolic and reproductive function in RED-S. The current findings are consistent with previous literature highlighting the utility of endocrine biomarkers as sensitive indicators of physiological recovery in RED-S. Increases in serum leptin, an adipocyte-derived hormone that signals energy sufficiency to the hypothalamus, are closely associated with improvements in reproductive function^57,58^. Similarly, elevations in triiodothyronine (TT3), a well-established marker of metabolic adaptation, reflect reversal of energy-conserving mechanisms and restoration of metabolic homeostasis^17^. Additional improvements in cortisol, oestradiol and LH pulsatility further support the reactivation of the hypothalamic–pituitary–gonadal (HPG) axis^19,52^. Given the low certainty of current evidence, future research must focus on high-quality, standardised studies to validate endocrine biomarkers as reliable indicators of RED-S recovery. Markers like leptin, TT3, oestradiol, and LH pulsatility show promise but are limited by inconsistent measurement, variability, and unclear clinical thresholds ^17,52^. To improve utility, studies should standardise biomarker timing, consider menstrual cycle, diurnal variation, and assay methods, and establish validated cut-offs. Longitudinal research linking hormonal changes to clinical outcomes such as menstrual resumption and bone health is needed. Importantly, biomarkers should be combined with clinical assessments, symptom tracking, and performance measures for a comprehensive evaluation^16^. Rigorous validation of these markers is essential to enhance RED-S diagnosis, monitoring, and management.

### Pharmacological Interventions

#### Bone mineral density

BMD outcomes in RED-S interventions were inconsistent across studies, with some evidence suggesting hormonal therapies may support bone health when nutritional recovery alone is inadequate.

These findings align with existing evidence suggesting that while hormonal therapies may offer some benefit to BMD^55^, their effectiveness remains inconsistent. For instance, a review by Indirli et al.,^59^ reported that hormone therapies using oestrogen or leptin showed limited impact on bone metabolism in women with FHA This variability is likely since such treatments do not address the underlying cause of RED-S: chronic low EA. Pharmacological interventions may alleviate certain symptoms but risk masking the broader physiological dysfunction if used in isolation^17,60^. As such, current evidence supports their use only as adjuncts in cases where nutritional rehabilitation and restoration of EA, the foundation of RED-S treatment, have not been sufficient, or where bone health is severely compromised^61^.

#### Menstrual function recovery

Although pharmacological interventions have shown promise in supporting menstrual recovery in RED-S, current evidence, particularly from non-pharmacological studies, suggests that nutritional and non-pharmacological strategies may offer equally, if not more, effective outcomes, albeit from a smaller and methodologically limited evidence base.

Evidence supporting the current findings aligns with established physiological mechanisms indicating that hormonal therapy in RED-S may obscure true recovery. Combined oral contraceptives (COCs) and other exogenous hormone regimens induce withdrawal bleeding through artificial endometrial shedding, without restoring endogenous ovulatory cycles or HPG axis function^62,63^. As such, withdrawal bleeding can be misinterpreted as menstrual recovery, potentially delaying appropriate treatment interventions targeting LEA. This masking effect is well-documented and underscores the need for more accurate markers of reproductive recovery. Objective indicators such as serum progesterone levels, LH pulsatility, or basal body temperature tracking are recommended to assess ovulatory function and HPG axis restoration^64^. Given these considerations, hormonal therapies should be used with caution and only as adjuncts to primary nutritional and non-pharmacological strategies that address the underlying energy deficiency central to RED-S.

#### Hormonal profiles

Hormonal interventions demonstrated significant and clinically meaningful effects on oestrogen and progesterone levels, with oestradiol notably higher in treatment groups compared to those receiving oral contraceptives. But certainty of evidence is mixed.

Ackerman et al.^16^ demonstrated that improvements in BMD can occur without endogenous hormonal normalization, as menstrual function did not resume during treatment with transdermal oestrogen. This indicates that symptom recovery, such as bone health improvement, may happen independently of menstrual and hormonal recovery when exogenous hormones are administered. Interpreting endocrine responses in this context is complex because exogenous hormones, like transdermal oestradiol or COC’s, suppress endogenous hormone production via negative feedback on the HPG axis^16,65^. This suppression can create misleading hormonal profiles that suggest normalization without true recovery of natural reproductive function, such as ovulation and menstrual cycles^65^. Therefore, clinical improvements in outcomes may reflect the direct effects of hormone therapy rather than restoration of endogenous endocrine function^16^. These complexities highlight the need for comprehensive assessment of RED-S recovery that goes beyond hormone levels to include clinical symptoms and functional reproductive status^17^.

#### Bone turnover markers

Analysis of bone turnover markers, specifically P1NP and IGF-1, showed statistically and clinically significant changes indicative of meaningful effects on bone metabolism, supported by high-certainty evidence; however, findings are limited by being based on a single study, highlighting the need for further research to validate their utility in monitoring bone health in athletes with RED-S.

These findings are supported by literature recognizing bone turnover markers such as P1NP and IGF-1 as sensitive indicators of bone metabolism^66^. However, their clinical utility is challenged by variability in assay methods, biological fluctuations, and influences from nutrition, exercise, and hormonal status^67^. The limited number of studies reporting these markers, alongside inconsistent measurement protocols and definitions, further complicates cross-study comparisons^68^. Nevertheless, bone turnover markers provide valuable early insight into bone remodelling processes that may occur before detectable changes in BMD, emphasizing the importance of standardized assessment methods and additional research to fully establish their role in monitoring bone health in RED-S.

### Limitations of the evidence

Most included studies forming the evidence were rated as having moderate to high risk of bias, and the consistency of using a standard set of reporting outcome measures was poor. This limits the strength of the evidence base. Reporting on intervention fidelity, participant compliance, and follow-up was often incomplete or unclear, reducing confidence in the consistency and applicability of reported outcomes. These methodological limitations highlight the need for more robust, standardised research in this field. Another limitation is that the standard deviation of change used in the meta-analysis for the Dueck^29^ was estimated.

### Implications for practice / clinical implications

The current review mostly included females from sports and events that are categorised as distance and endurance related, most non-pharmacological evidence utilised interventions that changed or adapted dietary intake and the most common environment was a University laboratory or location.

- Effective management of RED-S should prioritise restoring EA through non-pharmacological interventions. The current evidence consistently utilises dietary approaches delivered by dietitians.
- Recovery should not be assessed by menstrual status or body composition alone; instead, clinicians should adopt a multidimensional approach that incorporates hormonally validated markers (e.g., leptin, TT3, LH, oestradiol) and behavioural changes to ensure accurate diagnosis, monitoring, and long-term management.
- Hormonal therapies in RED-S may provide benefits for bone health and symptom management but should be used cautiously and only when nutritional rehabilitation and energy restoration are insufficient, as they do not address the underlying cause and can mask true reproductive recovery.
- Pharmacological treatments like COCs may induce withdrawal bleeding without restoring ovulatory function, emphasizing the need to prioritise nutritional and non-pharmacological interventions alongside objective monitoring. Additionally, bone turnover markers such as P1NP and IGF-1 show potential as sensitive indicators of bone metabolism, though their clinical utility is currently constrained by variability and limited evidence, underscoring the need for standardized assessment and further research.

## Conclusion

This review has provided consistent evidence from non-pharmacological interventions, primarily dietary adaptations, demonstrating beneficial effects on the restoration of menses, EA, fat mass and body fat percentage. Hormonal changes and biomarkers are currently supported by more consistent evidence from pharmacological studies compared to non-pharmacological studies. Further research is needed.

## Supporting information

https://bham-my.sharepoint.com/:w:/r/personal/a_a_soundy_bham_ac_uk/_layouts/15/Doc.aspx?sourcedoc=%7BC6335DC9-1980-4E93-9C42-F0A9BBA06AB2%7D&file=Sup

## Data Availability

All data produced in the present study are contained in the manuscript and supplementary documents

