## Supplementary material for "Pharmacological vs non-pharmacological treatment in the management of Relative Energy Deficiency in Sport (RED-S): A systematic review and meta-analysis": https://bham-my.sharepoint.com/:w:/r/personal/a_a_soundy_bham_ac_uk/_layouts/15/Doc.aspx?sourcedoc=%7BC6335DC9-1980-4E93-9C42-F0A9BBA06AB2%7D&file=Sup

Supplementary file

1. Critical appraisal results
2. Certainty Assessment results

*Critical Appraisal*

The following risk of bias tools were used depending on study design: ROB 2 (for Randomised Control trials) (RCTs, n= 7/19, 37%), ROBINS-I V2 (for non-randomised interventional studies, n= 11, 58%), and the JBI checklist (for case reports, n=1 , 5%).

Overall, RCTs demonstrated low (n= 3/7, 43%) to moderate (n= 4/7, 57%) risk of bias, with most (n= 5/7, 71%) studies showing low risk across all key domains. However, two RCTs (Michopoulos et al., 2013; Dadgostar et al., 2018) had "some concerns" particularly in randomisation and outcome reporting. In contrast, all non-randomised studies were rated as having serious risk due to the influence of confounding variables (e.g. baseline differences of energy availability, variation in training load, duration of menstrual dysfunction, age). Additional concerns were noted in non-RCTs in domains such as intervention classification (serious risk (n= 1/11, 9%), moderate risk (n= 10/11, 91%)), outcome measurement (n= 10/11, 91%), and missing data (n= 6/11, 55%), typically rated at moderate risk. The single case report by Mallinson et al. (2013) was methodologically sound across most JBI criteria but lacked reporting on adverse or unanticipated events.

| **ROB 2** | | | | | | |
|---|---|---|---|---|---|---|
| | bias arising from the randomisation process | bias due to deviations from intended interventions | bias due to missing outcome data | bias in measurement of the outcome | bias in selection of reported results | overall ranking |
| Randomised controlled trial of the effects of increased energy intake on menstrual recovery in exercising women with menstrual disturbances: the 'REFUEL' study. De Souza et al (2021) | Low | Low | Moderate | Low | Low | low |

| | | | | | | |
|---|---|---|---|---|---|---|
| Treatment of reduced bone mineral density in athletic amenorrhea: a pilot study<br>Gibson et al (1999) | Low | Low-Moderate | Moderate | Low | Low | moderate |
| Persistent osteopenia in ballet dancers with amenorrhea and delayed menarche despite hormone therapy: a longitudinal study<br>Warren et al (2003) | Low | Low | Some concerns | Low | Low | low |
| Neuroendocrine recovery initiated by cognitive behavioral therapy in women with functional hypothalamic amenorrhea: a randomized, controlled trial<br>Michopoulos et al (2013) | Some concerns | Some concerns | Low | Low | Some concerns | moderate |
| Oestrogen replacement improves bone mineral density in oligo-amenorrhoeic athletes: a randomised clinical trial<br>Ackerman et al. (2019) | Low | Some concerns | Low | Low | Some concerns | moderate |

| | | | | | | |
|---|---|---|---|---|---|---|
| Bone mineral density in response to increased energy intake in exercising women with oligomenorrhea/amenorrhea: the REFUEL randomized controlled trial De Souza et al. (2022) | Low | Some concerns | Low | Low | Low | low |
| The effect of hormone therapy on bone mineral density and cardiovascular factors among Iranian female athletes with amenorrhea/oligomenorrhea: A randomised clinical trial Dadgostar et al (2018) | Some concerns | Some concerns | Low | Low | Some concerns | moderate |

| ROBINS I V2 | | | | | | | |
|---|---|---|---|---|---|---|---|
| | risk of bias due to confounding | risk of bias in classification of intervention | risk of bias in selection into the study | risk of bias due to deviations from intended intervention | risk of bias due to missing data | risk of bias arising from measurement of the outcome | risk of bias in selection of the reported results |
| Energy and Nutrient Status of Amenorrheic Athletes Participating in a Diet and Exercise Training Intervention Programme Kopp-Woodroffe et al. (1999) | serious risk | moderate risk | low risk | moderate risk | low risk | moderate risk | moderate risk |
| Participant evaluations of the FUEL intervention designed for female endurance athletes at risk of REDs: A mixed methods approach. Solstad et al. (2025) | serious risk | moderate risk | low risk | low risk | moderate risk | moderate risk | moderate risk |
| Healthy Runner Project: a 7-year, multisite nutrition education intervention to reduce bone stress injury incidence in collegiate distance runners Fredericson et al (2023) | serious risk | moderate risk | low risk | moderate risk | low risk | low risk | moderate risk |

| | | | | | | | |
|---|---|---|---|---|---|---|---|
| Effects of a 16-week digital intervention on sports nutrition knowledge and behavior in female endurance athletes with risk of relative energy deficiency in sport (REDs)<br>Fahrenholtz et al (2023) | serious risk | moderate risk | low risk | low risk | moderate risk | moderate risk | low risk |
| Dietary intervention restored menses in female athletes with exercise-associated menstrual dysfunction with limited impact on bone and muscle health Cialdella-Kam et al (2014) | serious risk | serious risk | low risk | moderate risk | low risk | moderate risk | moderate risk |
| Active women before/after an intervention designed to restore menstrual function: resting metabolic rate and comparison of four methods to quantify energy expenditure and energy availability<br>Guebels et al (2014) | serious risk | moderate risk | low risk | moderate risk | low risk | moderate risk | moderate risk |
| Nine–month nutritional intervention improves restoration of menses in young female athletes and ballet dancers<br>Lagowska et al (2014) | serious risk | moderate risk | low risk | moderate risk | moderate risk | moderate risk | moderate risk |

| | | | | | | | |
|---|---|---|---|---|---|---|---|
| Effects of dietary intervention in young female athletes with menstrual disorders<br>Łagowska et al (2014) | serious risk | moderate risk | low risk | moderate risk | moderate risk | moderate risk | moderate risk |
| Treatment of athletic amenorrhea with a diet and training intervention program<br>Dueck et al., (1996) | serious risk | moderate risk | low risk | moderate risk | low risk | moderate risk | moderate risk |
| Restoration of menses with nonpharmacologic therapy in college athletes with menstrual disturbances: a 5-year retrospective study<br>Aredns et al (2012) | serious risk | moderate risk | moderate risk | serious risk | serious risk | moderate risk | moderate risk |
| Relative Energy Deficiency in Sport—Multidisciplinary Treatment in Clinical Practice<br>Meyer et al (2025) | serious risk | moderate risk | moderate risk | serious risk | moderate-serious risk | moderate risk | moderate risk |

| | JBI | | | | | | | |
|---|---|---|---|---|---|---|---|---|
| | Were patient's demographic characteristics clearly described? | Was the patient's history clearly described and presented as a timeline? | Was the current clinical condition of the patient on presentation clearly described? | Were diagnostic tests or assessment methods and the results clearly described? | Was the intervention(s) or treatment procedure(s) clearly described? | Was the post-intervention clinical condition clearly described? | Were adverse events (harms) or unanticipated events identified and described? | Does th report p takeaw lessons |
| A case report of recovery of menstrual function following a nutritional intervention in two exercising women with amenorrhea of varying duration Mallinson et al. (2013) | yes | yes | yes | yes | yes | yes | no | yes |

| | **Menstrual function recovery** | | | **Energy availability** | | | **Body composition** | | | **Biomarkers** | |
|---|---|---|---|---|---|---|---|---|---|---|---|
| | *GRADE assessment of certainty* | *Reasons for downgrade (-) or upgrade (+)* | | *GRADE assessment of certainty* | *Reasons for downgrade or upgrade* | | *GRADE assessment of certainty* | *Reasons for downgrade or upgrade* | | *GRADE assessment of certainty* | *Reasons for downgrade or upgrade* |
| Kopp-Woodroffe et al. (1999) | | | | Low | risk of bias (-1), imprecision (-1), publication bias (-1), large effect size (+1) | | | | | Low | risk of bias (-1), imprecision (-1), publication bias (-1), moderate effect size (+1) |
| Solstad et al. (2025) | | | | | | | | | | | |
| Mallinson et al. (2013) | Very low | risk of bias(-1), inconsistency (-1), imprecision (-1), publication bias (-1) | | | | | Low | risk of bias(-1), imprecision (-1), publication bias (-1), very large effect size (+1) | | Low | risk of bias (-1), imprecision (-1), publication bias (-1), large effect size (+1) |
| De Souza et al (2021) | High | Imprecision (-1), large effect (+1), dose-reponse (+1) | | | | | Very high | large effect size (+1) | | Very high | |
| Meyer et al (2025) | Low | risk of bias (-1), indirectness (-1), imprecision (-1), publication bias (-1), very large effect size (+1) | | | | | Low | risk of bias(-1), imprecision (-1), publication bias (-1), large effect size (+1) | | | |

| | | | | | | | | | | | |
|---|---|---|---|---|---|---|---|---|---|---|---|
| Fredericson et al (2023) | | | | | | | | | | | |
| Fahrenholtz et al (2023) | | | | | | | | | | | |
| Cialdella-Kam et al (2014) | Low | risk of bias (-1), imprecision (-1), publication bias (-1), large effect size (+1) | | Low | risk of bias (-1), imprecision (-1), publication bias (-1), moderate effect size (+1) | | | | | Low | risk of bias (-1), imprecision (-1), publication bias (-1), moderate effect size (+1) |
| Guebels et al (2014) | | | | Low | risk of bias (-1), imprecision (-1), publication bias (-1), large effect size (+1) | | | | | | |
| Lagowska et al (2014) | Low | risk of bias (-1), imprecision (-1), publication bias (-1), large effect size (+1) | | Low | risk of bias (-1), imprecision (-1), publication bias (-1), moderate effect size (+1) | | | | | Low | risk of bias (-1), imprecision (-1), publication bias (-1), moderate (+1) |

| | | | | | | | | | | | |
|---|---|---|---|---|---|---|---|---|---|---|---|
| Gibson et al (1999) | | | | | | | | | | Very high | moderate effect size (+1) |
| Warren et al (2003) | | | | L ow | risk of bias (-1), imprecision (-1), publication bias (-1), large effect size (+1) | | Low | risk of bias(-1), imprecision (-1), publication bias (-1), moderate effect size (+1) | | Low | risk of bias (-1), imprecision (-1), publication bias (-1), moderate effect (+1) |
| Michopoulos et al (2013) | High | | | | | | Very high | moderate effect size (+1) | | Very high | moderate effect size (+1) |
| Łagowska et al (2014) | Low | risk of bias (-1), imprecision (-1), publication bias (-1), large effect size (+1) | | | | | Low | risk of bias(-1), imprecision (-1), publication bias (-1), moderate effect size (+1) | | Low | risk of bias (-1), imprecision (-1), publication bias (-1), moderate effect size (+1) |
| Ackerman et al. (2019) | Low | risk of bias (-1), imprecision (-1), publication bias (-1), very large effect size (+1) | | | | | Low | risk of bias(-1), imprecision (-1), publication bias (-1), moderate effect size (+1) | | | |

| | | | | | | | | | | | |
|---|---|---|---|---|---|---|---|---|---|---|---|
| **Author** | **Bone Mineral Density (BMD) (via DXA scan)** | | | **Hormonal profiles** | | | **Menstrual function recovery** | | | **Bone turnover markers** | |
| | GRADE assessment of certainty | Reasons for downgrade (-) or upgrade (+) | | GRADE assessment of certainty | Reasons for downgrade (-) or upgrade (+) | | GRADE assessment of certainty | Reasons for downgrade (-) or upgrade (+) | | GRADE assessment of certainty | Reasons for downgrade (-) or upgrade (+) |
| De Souza et al. (2022) | Low | risk of bias (-1), imprecision (-1), publication bias (-1), large effect size (+1) | | Low | risk of bias (-1), imprecision (-1), publication bias (-1), large effect size (+1) | | Low | risk of bias (-1), imprecision (-1), publication bias (-1), large effect size (+1) | | | |
| Dueck et al., (1996) | Very low | risk of bias (-1), imprecision (-1), publication bias (-1) | | Very low | risk of bias (-1), imprecision (-1), publication bias (-1) | | Very low | risk of bias (-1), imprecision (-1), publication bias (-1) | | | |
| Aredns et al (2012) | Very high | large effect size (+1) | | Very high | moderate effect size (+1) | | High | | | High | |
| Dadgostar et al (2018) | High | | | | | | | | | | |
